# Assessment of Socio-Demographic factors associated with the Utilization of Insecticide-Treated Nets (ITNs) Among Children Under-5 years old and Pregnant Women in Nigeria: A secondary analysis of NMIS data

**DOI:** 10.1101/2023.11.30.23299264

**Authors:** Emmanuel Babagbotemi Omole, Olachi Sandra Ndukwe

## Abstract

Malaria remains a persistent global health challenge, particularly in sub-Saharan Africa, with Nigeria shouldering a disproportionate burden. Despite extensive interventions, insecticide-treated nets (ITNs) have emerged as a cost-effective tool in the World Health Organization’s malaria control strategy. Utilizing data from the 2021 Nigeria Malaria Indicator Survey, this study explores socio-demographic determinants influencing ITN utilization among specific cohorts—children under-5 years and pregnant women. Findings indicate that 56% of Nigerian households possessed at least one ITN, with significant utilization observed among rural households, households from the North West and North East geopolitical zones, and households in the second and lowest wealth quintiles (*p* <0.05). Only 41.2% of children under-5 years old slept under an ITN on the night before the survey, underscoring notable coverage gaps. Statistical analyses reveal significant associations (*p* < 0.05) between ITN usage and variables such as age, residence, geopolitical zone, and wealth quintile at both bivariate and multivariate analytical levels. ITN usage decreases with increasing child age and household wealth quintile. For pregnant women, almost half (50%) in all households slept under an ITN the night before the survey. At both bivariate and multivariate analytical levels, significant associations (*p* < 0.05) were observed between ITN usage among pregnant women and variables such as geopolitical zone and household wealth quintile. Additionally, at the multivariate level, ITN utilization decreases with increasing educational level. Our study reveals the dynamic nature of ITN usage patterns, necessitating ongoing monitoring and adaptive strategies to address regional and socioeconomic differentials, while sustaining awareness initiatives to meet the targets set by the Nigeria National Malaria Strategic Plan.

## 1. Introduction

Malaria persists as a major global health burden which continues to worsen, with 247 million cases and 619,000 related deaths reported globally in 2021 [1]. Malaria remains endemic in most sub-Saharan African countries, constituting 85% of global cases and accounting for almost 93% of all malaria-related deaths worldwide. Unfortunately, over three-quarters of these mortalities are children under the age of 5 [2, 3].

Among these sub-Saharan African countries, Nigeria carries the highest burden, representing nearly 27% of the global malaria cases [2]. According to the World Health Organization (WHO), Nigeria reported an estimated 68 million cases and 194,000 deaths from malaria in 2021, making it a major public health concern [4]. The economic toll is equally substantial, with Nigeria losing about 646 billion Naira ($1.1 billion USD) annually due to malaria-related cases [5].

In the global battle against malaria, a myriad of interventions has been employed. These interventions constitute the WHO-recommended package to prevent and reduce morbidity and mortality. They include vector control through in-door residual spraying and insecticide-treated nets, seasonal chemoprevention, rapid diagnostic testing, and treatment with artemisinin-based combination therapies [6,7,8]. More recently, the groundbreaking utilization of malaria vaccines has been incorporated into this comprehensive approach [7,8]. Despite their effectiveness, the implementation of these interventions entails substantial costs [9,10]. Notably, among these strategies, the use of insecticide-treated nets (ITNs) has emerged as the most cost-effective, contributing significantly to averting over 50% of malaria cases and reducing child mortality by 27% [11,12]. Over the last 2 decades, ITNs have contributed significantly to the progress seen in reducing malaria cases worldwide and drove most of the declines in malaria seen from 2005–2015, especially in moderate-to-high transmission areas [12].

The use of ITNs remains a crucial component of the WHO’s strategy to combat the transmission of malaria in malaria-endemic regions, especially sub-Saharan Africa [7,8]. The distribution of ITNs in this region has seen a significant increase in an effort to protect every household at risk of malaria transmission, with a particular focus on pregnant women and children under the age of five [13, 14,15].

The Abuja Declaration, signed in 2000, marked a pivotal moment in the fight against malaria, with 44 malaria-endemic countries in Africa committing to halving malaria-attributable mortality by 2010 and protecting 60% of pregnant women and children under-5 years with measures such as ITNs by 2005 [16]. The effectiveness of ITNs, demonstrated by reducing clinical malaria by over 50% and decreasing all-cause mortality in young children by 15-30% when overall population usage exceeds 70%, underscores their critical role in achieving these ambitious goals [17,18,19]. As the Roll Back Malaria Partnership evolved, so did the strategies and targets [8]. The Nigeria National Malaria Strategic Plan (NMSP) builds on these efforts, aiming to improve access and utilization of vector control interventions to 80% of the target population by 2025 [20]. The NMSP includes priorities such as reducing malaria-related mortality, decreasing malaria parasite prevalence in children under-5 years, and increasing ownership and use of ITNs and long-lasting insecticidal nets (LLINs) [20].

Over the past decade, Nigeria has made significant progress in improving the utilization of ITNs among children under-5 years old and pregnant women [6,21]. The implementation of various malaria control strategies, including mass distribution campaigns, antenatal care programs, and community-based interventions, has contributed to increased ITN usage [6]. Despite the overall progress, disparities in ITN utilization still persist. Factors contributing to these disparities include differences in access to healthcare services, socioeconomic conditions, level of maternal education, household wealth, and awareness of the importance of ITNs [21, 22]. This study was aimed at determining some demographic factors associated with the use of ITNs among children under 5-years and pregnant women in Nigeria based on the recent 2021 Nigeria Malaria Indicator Survey (NMIS).

## 2. Materials and Methods

### Study design

This study was a secondary analysis based on data drawn from the Nigeria Malaria Indicator Survey (NMIS) 2021 [21]. The 2021 NMIS marks the third iteration of malaria indicator surveys conducted in Nigeria, following the inaugural survey in 2010 and the subsequent one in 2015. The survey collected data on household ownership of an ITN, and number of children who slept under an ITN on the night preceding the survey. The survey defined an ITN as a factory treated net not requiring any further treatment. The data collection process also encompassed sociodemographic and economic characteristics of the household and child, such as the child’s age, sex, residence, geopolitical zone, and household wealth quintile.

### Study population

The survey was conducted in 13,727 households and a total of 14,476 women aged 15-49 years from the selected households were successfully interviewed in the survey.

### Data analysis

Data analysis was performed in two phases. In the first phase, data abstracted from the NMIS were uploaded into R version 4.3.2 statistical software and descriptive analysis was carried out to describe the selected socio-demographic characteristics. Furthermore, an association between the use of ITNs and socio-demographic variables was assessed with the chi-square test to explore the individual effect of socio-demographic factors on ITN usage. In the second phase, odd ratios (OR) and the respective 95% confidence interval (CI) were calculated to determine the strength of association.

## 3. Results

### Household Ownership of ITNs

The initial analysis of the data covers the association between ITN access and various socio-demographic variables. As shown in Table 1, 56% of Nigerian households possessed at least one ITN. If it is assumed that one ITN can be shared by two people who stayed in the household on the night preceding the survey, 25% of households own enough ITNs to cover all household members. This translates to an average of 1.3 ITNs per household.

**Table 1.** Households in possession of at least one ITN.^Ω^.

| **Table 1. Households in possession of at least one ITN.<sup>Ω</sup>** |  |  |  |
| --- | --- | --- | --- |
| **Characteristic** | **No of Households *n*<sup>Δ</sup>** | **Households with ITNs *n*<sup>Δ</sup> (%)** | **p-value<sup>†</sup>** |
| **Residence** |  |  | 0.005* |
| Urban | 4,546 | 2400 (52.8) |  |
| Rural | 9,181 | 5279 (57.5) |  |
| **Geopolitical Zone** |  |  | <0.001* |
| North Central | 2,210 | 1103 (49.9) |  |
| North East | 2,089 | 1504 (72.0) |  |
| North West | 3,629 | 2751 (75.8) |  |
| South East | 1,356 | 500 (36.9) |  |
| South South | 2,037 | 801 (39.3) |  |
| South West | 2,406 | 1023 (42.5) |  |
| **Wealth quintile** |  |  | <0.001* |
| Lowest | 2,219 | 1420 (64.0) |  |
| Second | 2,365 | 1615 (68.3) |  |
| Middle | 2,707 | 1621 (59.9) |  |
| Fourth | 3,018 | 1536 (50.9) |  |
| Highest | 3,418 | 1487 (43.5) |  |
| **Total** | **13,727** | **7687 (56.0)** |  |

<sup>Ω</sup>Table is based on households with at least one ITN per two persons who stayed in the household the night before the survey.
<sup>Δ</sup>*n* may vary by characteristic due to missing data <sup>†</sup>p-value calculated using chi-squared test.

Subsequent analysis using the chi-square test, as presented in Table 1, indicated a significant association between ITN ownership in Nigeria and factors such as place of residence, geopolitical zone, and household wealth quintile. Specifically, rural households (*p* = 0.005), households from the North West and North East geopolitical zones (*p* <0.001), and households in the second and lowest wealth quintiles (*p* <0.001) possessed a significantly higher number of ITNs compared to their respective counterparts [Table 1].

### ITN Usage Among Children Under 5 Years Old

An average of only 41.4% of children under 5 years old in all households surveyed slept under an ITN on the night before the survey [Table 2]. The likelihood of a child sleeping under an ITN was found to be significantly associated with the child’s age (*p* = 0.007), residence (*p* = 0.001), geopolitical zone (*p* <0.001), and the wealth quintile of the household (*p* <0.001), and not significantly associated with sex (p=0.665). The use of ITNs decreased with increasing child age and household wealth quintile. Children under 12 months old (OR 1.21, 95% CI 1.09-1.33, *p* <0.001) were 1.2 times more likely to sleep under an ITN compared to 4-year-olds (48–59 months old). Additionally, children in rural areas (OR 1.14, 95% CI 1.06-1.22, *p* =0.001) had a 1.14-fold likelihood of sleeping under an ITN compared to their urban counterparts. Geopolitical zone also significantly impacts the usage of ITNs, with children in the North West (OR 2.46, 95% CI 2.14-2.82, *p* <0.001) and North East (OR 2.28, 95% CI 1.98-2.65, *p* <0.001) regions being twice as likely to sleep under an ITN compared to those from other geopolitical zones. Lastly, children from lower wealth quintiles—lowest (OR 1.58, 95% CI 1.41-1.77, *p* <0.001), second (OR 1.69, 95% CI 1.51-1.89, *p* <0.001), and middle (OR 1.53, 95% CI 1.37-1.72, *p* <0.001)—were 1.5 to 1.6 times more likely to sleep under an ITN than those in the highest wealth quintile.

**Table 2.**
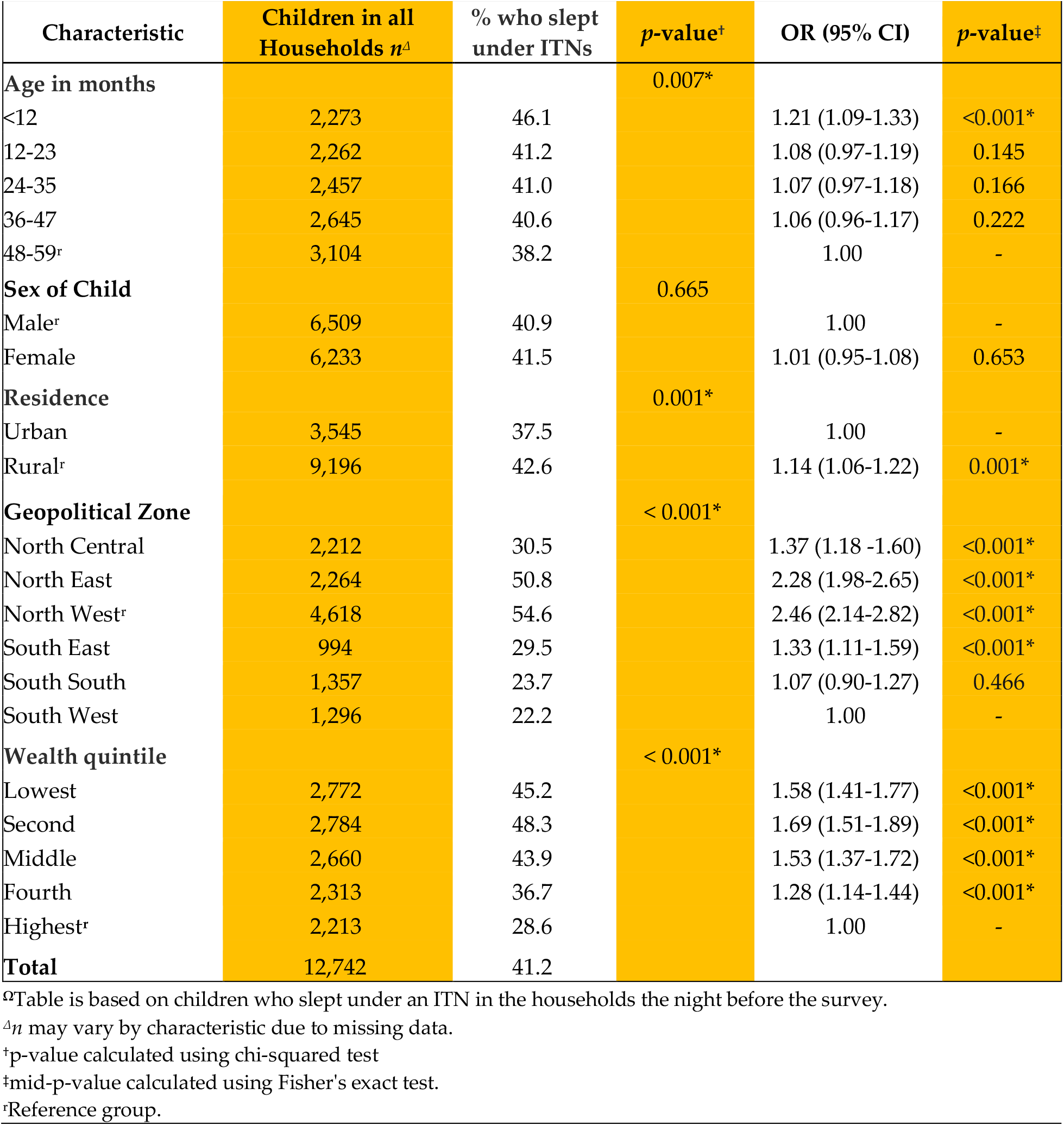
Children who slept under an ITN in households with at least one ITNs.^Ω^.

### ITN Usage Among Pregnant Women

Almost one-half of pregnant women in all households slept under an ITN the night before the survey [Table 3]. As with our data for children under the age of 5, various socio-dem-graphic variables significantly impact the usage of ITNs. Pregnant women from the North East (OR 2.86, 95% CI 1.79-4.75, *p* <0.001) and North West (OR 2.61, 95% CI 1.67-4.24, *p* <0.001) are twice as likely to sleep under an ITN compared to those from other geopolitical zones. Interestingly, we found that ITN usage was less common amongst more educated women, as women with no (1.46, 95% CI 1.00-2.18, *p*=0.048) or primary (1.57, 95% CI 1.02-2.44, *p*=0.042) education were roughly 1.5 times more likely to use an ITN compared to women who received beyond primary education. Similar to Table 2, the use of ITNs increases with decreasing household wealth status, as pregnant women from lower wealth quintiles—lowest (OR 1.82, 95% CI 1.27-2.63, *p* <0.001), second (OR 1.82,, 95% CI 1.28-2.61, *p* <0.001), middle (OR 1.90, 95% CI 1.34-2.73, *p* <0.001), and fourth (OR 1.47,, 95% CI 1.01-2.15, *p* =0.042)—were 1.4 to almost 2 times more likely to sleep under an ITN than those from the highest wealth quintile.

**Table 3.** Pregnant women who slept under an ITN in all households.^Ω^.

| Characteristic | Pregnant women in all households $n^A$ | % who slept under ITNs | $p$-value<sup>†</sup> | OR (95% CI) | $p$-value<sup>‡</sup> |
| --- | --- | --- | --- | --- | --- |
| **Residence** |  |  | 0.158 |  |  |
| Urban | 357 | 157 (44.1) |  | 1.00 | - |
| Rural | 963 | 498 (51.7) |  | 1.18 (0.95-1.46) | 0.142 |
| **Geopolitical Zone** |  |  | <0.001* |  |  |
| North Central | 184 | 65 (35.1) |  | 1.55 (0.93 - 2.67) | 0.095 |
| North East | 247 | 161 (65.0) |  | 2.86 (1.79-4.75) | < 0.001* |
| North West | 615 | 366 (59.5) |  | 2.61 (1.67-4.24) | < 0.001* |
| South East | 73 | 20 (27.4) |  | 1.21 (0.62-2.36) | 0.577 |
| South South | 94 | 19 (20.6) |  | 0.89 (0.45-1.74) | 0.743 |
| South West | 106 | 24 (22.8) |  | 1.00 | - |
| **Education** |  |  | 0.112 |  |  |
| No education | 614 | 325 (52.9) |  | 1.46 (1.00-2.18) | 0.048* |
| Primary | 175 | 99 (56.8) |  | 1.57 (1.02-2.44) | 0.042* |
| Secondary | 420 | 191 (45.4) |  | 1.26 (0.85-1.90) | 0.255 |
| More than secondary | 111 | 40 (36.2) |  | 1.00 | - |
| **Wealth quintile** |  |  | 0.003* |  |  |
| Lowest | 273 | 149 (54.6) |  | 1.82 (1.27-2.63) | < 0.001* |
| Second | 308 | 168 (54.6) |  | 1.82 (1.28-2.61) | < 0.001* |
| Middle | 298 | 170 (57.0) |  | 1.90 (1.34-2.73) | < 0.001* |
| Fourth | 257 | 113 (44.0) |  | 1.47 (1.01-2.15) | 0.042* |
| Highest | 184 | 55 (29.9) |  | 1.00 | - |
| **Total** | 1,320 | 655 (49.6) |  |  |  |

<sup>Ω</sup>Table is based on pregnant women age 15 – 49 who slept under an ITN in the households the night before the survey.
<sup>A</sup> $n$ may vary by characteristic due to missing data.
<sup>†</sup> $p$ -value calculated using chi-squared test.
<sup>‡</sup>mid- $p$ -value calculated using Fisher's exact test.
<sup>\*</sup>Referent group.

## 4. Discussion

In the past decade, Nigeria has made considerable progress in improving the use of ITNs among children under-5 years old and pregnant women. This study aimed to assess the association between socio-demographic factors and the use of Insecticide-Treated Nets (ITNs) in children under-5 years old and pregnant women in Nigeria, using nationally representative data from the 2021 Nigeria Malaria Indicator Survey (NMIS). The percentage of households possessing at least one ITN experienced a substantial rise from 8% in 2008 to 69% in 2015, before subsequently declining to 56% in 2021, and this decline can be attributed to the COVID-19 pandemic thus overshadowing some of the previously recorded gains [21, 23]. However, a quick snapshot review of comparative data from other Sub-Saharan African countries in 2021 showed higher ownership rates in Niger (96%), Mali (91%), Burkina Faso (93%), Cote d’Ivoire (72%), Madagascar (69%), and Guinea (63%) [24]. The substantial rise in ownership within households over the past decade can be attributed to the more assertive implementation of various reported malaria control strategies, including mass distribution campaigns, antenatal care (ANC) initiatives, and community-based interventions, fostering increased ITN utilization [25]. According to the NMIS 2021 report [21], over three-quarters of ITNs in Nigerian households were acquired through mass distribution campaigns, with other sources including markets, ANC visits, immunization visits, and government or private health facilities. In this study, factors such as place of residence, geopolitical zone, and household wealth quintile were significantly associated with ITN ownership in Nigeria. Notably, household ownership of ITNs is higher in rural areas (58%) compared to urban areas (53%). Additionally, regional disparities are evident, with household ownership of ITNs being highest in the North West (76%) and lowest in the South East (37%). Also noteworthy is that households in the second and lowest wealth quintiles possessed a significantly higher number of ITNs (68% and 64%, respectively) compared to the remaining quintiles, especially the highest wealth quintile (44%). These findings tend to reflect the successful reach of ITN distribution programs to rural communities and positive aftermath of several strategies to improve ownership of ITNs targeted at these population groups that have had low ITN coverage in the past [22, 26].

The findings of this study also provide valuable insights into the factors influencing ITNs utilization among specific population groups, namely children under-5 years old and pregnant women. Assessing ITN usage among children under-5 years reveals disparities between households. In all households, an overage of only 41.4% of children under-5 years slept under an ITN, exposing a notable gap in optimal coverage. At both bivariate and multivariate levels, ITN usage was significantly associated with age, residence, geopolitical zone, and wealth quintile. Younger children and those in rural areas showed higher likelihoods of sleeping under ITNs. The North West and North East regions stood out with a twofold likelihood of ITN usage, underlining regional variations. Lower wealth quintiles exhibited a 1.5 to 1.6 times higher likelihood of ITN usage than the highest quintile. Other similar studies report that children under 12 months are still being breastfed by their mothers who are more likely to share a bed covered with ITN with them, and this tends to diminish with increasing age of the children [27,28]. Increase in ITN usage observed in financially disadvantaged households may be attributed to the economic conditions, wherein individuals in these households tend to share beds, as opposed to wealthier households that have the luxury of more separate sleeping arrangements. Another study from Nigeria reported that ITN utilization increases when children share beds with other family members [29].

In the present study, ITN usage among pregnant women varies between all households. This variation offers insights into the impact of ITN availability within a household. In all households, about half of pregnant women (50%) used an ITN, with a significant association found at the bivariate analytical level, linking this usage to geopolitical zone and household wealth quintile. Pregnant women from the North East and North West regions as well as those from middle, second and lowest quintiles utilizes ITN more. At the multivariate level, pregnant women from the North East and North West regions had twice the odds of using an ITN compared to other zones, while the odds decreased with increasing educational level and increasing household wealth status. A similar study done in Nigeria also reports higher ITN ownerships and usage among pregnant women in the North East and North West [30]. These two geopolitical zones tend to be more rural in nature, with extreme poverty and illiteracy rates than the urbanized South West. Thus, the increased utilization reported in this present study may reflect the success of targeted interventions through massive community-level distribution of ITNs at antenatal care facilities in these areas.

Our study reveals the dynamic nature of ITN usage patterns which should necessitate ongoing monitoring and adaptive strategies. Regular assessments of demographic and socio-economic trends can inform timely adjustments to intervention programs, ensuring their relevance and effectiveness over time. Additionally, continuous efforts are essential to bridge the gap in ITN usage among specific population groups and ensure comprehensive malaria prevention in diverse household settings.

## 5. Conclusions

In conclusion, while strides have been made in improving ITN utilization among children under 5 and pregnant women, it is far from the target of 80% set for both indicators in the NMSP. Addressing identified regional disparities, socioeconomic factors, and maintaining awareness initiatives are pivotal for achieving universal and equitable ITN coverage. These interventions should include increasing access to healthcare services, expanding ITN distribution programs, and strengthening health education initiatives. These inferences guide future interventions to ensure sustained progress in malaria prevention efforts across diverse populations in Nigeria.

## Data Availability

The data that support the findings of this research is accessible through the Demographic and Health Surveys (DHS) Program upon reasonable requests and approval from the DHS Program. The datasets analyzed in this research can be found in the DHS repository at https://dhsprogram.com/data/available-datasets.cfm. The codes for the data analysis used in this research are available upon request.

## Author Contributions

Conceptualization, E.B.O. and O.S.N.; methodology, E.B.O. and O.S.N.; data curation, E.B.O. and O.S.N.; software, E.B.O.; formal analysis, E.B.O. and O.S.N.; writing—original draft preparation, E.B.O. and O.S.N.; writing—review and editing, E.B.O. and O.S.N.; visualization, E.B.O. and O.S.N.; supervision, E.B.O.; project administration, E.B.O. and O.S.N.

## Funding

This research received no external funding.

## Acknowledgments

The authors would like to express their gratitude to the Demographic and Health Surveys (DHS) Program for access to the survey datasets used for this study and to James Richard Ellegate Jr for his guidance in R programming and revision of the manuscript materials.

## Conflicts of Interest

The authors declare no conflict of interest.

## Abbreviations

DHS: Demographic and Health Surveys
ITN: Insecticide-treated net
LLIN: Long-Lasting Insecticidal Net
NMIS: Nigeria Malaria Indicator Survey
NMSP: Nigeria National Malaria Strategic Plan
WHO: World Health Organization

## Notes

### Competing Interest Statement

The authors have declared no competing interest.

### Summary of Updates

Tables arrangement adjusted for clarity.

